# The Puppy Escape Narrative: Validation of an Openly Available Recall Task for MCI Detection

**DOI:** 10.64898/2026.07.01.26357019

**Authors:** Michael J Kleiman, Dierdre O’Shea, Katana Rader, Mirza Baig, Simone Camacho, Andres Salcedo, James E Galvin

**Affiliations:** Comprehensive Center for Brain Health, Department of Neurology, University of Miami Miller School of Medicine, Boca Raton, FL 33433

**Keywords:** narrative recall, mild cognitive impairment, neuropsychological assessment, Alzheimer’s disease, episodic memory, psychometric validation, cognitive screening

## Abstract

**Introduction:** Narrative recall is widely used to detect cognitive impairment, but dominant instruments carry proprietary restrictions. The Craft Story 21 (CS), the non-proprietary NACC UDS4 standard, is not available standalone. Here, we validate the freely available Puppy Escape (PE).

**Methods:** 346 participants (153 cognitively normal, 106 subjective cognitive impairment, 87 mild cognitive impairment) completed PE and CS. Analyses evaluated convergent and criterion validity, MCI-vs-control discrimination, and incremental validity.

**Results:** PE and CS converged (r=.43–.47) and were equivalent on 10/12 neuropsychological measures. PE Delayed discriminated MCI from controls (d=1.03; ROC-AUC equal to CS, DeLong p=.510) and added variance beyond CS (ΔR²=+.054, p<.001). Automated subscores revealed MCI deficits in location, action, and name content. PE-18 short form retained discrimination (d=1.02) with 18 items.

**Discussion:** PE matched CS across all validation domains and captured complementary diagnostic information. PE and PE-18 are available via online registration explicitly permitting industry-sponsored research and fee-for-service clinical use.

## 1. Background

Narrative recall has served as a cornerstone of neuropsychological assessment since Wechsler incorporated prose passage memory into the first standardized clinical memory battery [1], and paragraph-length story recall tasks remain as widely administered instruments for detecting cognitive impairment in aging populations. The clinical prominence of narrative recall rests on a direct neurobiological rationale: verbal episodic memory, the capacity to encode, consolidate, and subsequently retrieve a temporally organized account of experienced events, is among the earliest and most reliably disrupted cognitive domains in Alzheimer’s disease (AD) and its prodromal stage, mild cognitive impairment (MCI) [2].

Despite this longstanding clinical utility, the most widely validated narrative recall instruments carry access restrictions that increasingly conflict with the need for scalable, equitable cognitive screening. The widely-used Wechsler Memory Scale Logical Memory (WMS-LM) [3] and Alzheimer’s Disease Assessment Scale–Cognitive Subscale (ADAS-Cog) narrative subtest [4] carry per-subject licensing fees, kit-purchase requirements, or proprietary distribution restrictions. These licensing barriers effectively exclude community health centers, low-resource clinical settings, and international sites from deploying the most extensively normed narrative recall instruments, creating a widening tension between populations most in need of early detection and those with practical access to validated tools [5]. The Craft Story 21 (CS) was introduced into the National Alzheimer’s Coordinating Center (NACC) Uniform Data Set Version 3 (UDS-3) and then Version 4 (UDS-4) specifically to address this access problem, replacing the proprietary Logical Memory subtest as the standard narrative recall task across the National Institute on Aging Alzheimer Disease Research Center (ADRC) network [6,7]. CS has not fully resolved the access problem, however: it still requires developer permission for distribution outside the UDS-4 materials package, lacks a standardized automated scoring pathway, and largely remains embedded within the ADRC network, leaving deployment outside that ecosystem administratively dependent on the NACC apparatus rather than freely scalable to independent research sites, community clinics, or international settings.

Existing narrative recall instruments thus have no alternative option that simultaneously offers free availability for clinical and research use with no per-subject cost, validated manual and automated scoring pathways, and normative validation across the cognitively normal to MCI continuum [8–11]. The Puppy Escape (PE) narrative recall task was developed to address this gap: a paragraph-recall story designed for both pen-and-paper clinical deployment and computational scoring, freely available for clinical and research use, with a 57-item automated rubric mapping each recalled detail to one of six semantic categories and five narrative events and an 18-item manually-scorable short form (PE-18) derived from that rubric for clinical use without computational infrastructure. The PE narrative’s increased word count and reduced delay period of 15-minutes compared to previous instruments’ standard of 30-minutes enables easier and more rapid assessment at a lower resource cost.

This study reports a comprehensive validation of the Puppy Escape against the Craft Story 21. Six pre-specified objectives were evaluated: (1) convergent validity between PE and CS, hypothesizing moderate-to-strong correlations; (2) criterion validity against an established neuropsychological battery, hypothesizing statistical parity between the two stories; (3) CN-vs-MCI group discrimination, hypothesizing clinically meaningful PE effect sizes with DeLong comparison to CS; (4) incremental validity of PE beyond CS, hypothesizing significant incremental prediction given differences in narrative complexity and scoring; (5) dimensional profiling of MCI impairment across PE’s semantic category and narrative event subscores, hypothesizing selective impairment of content carrying the strongest contextual-binding load; and (6) derivation and validation of a parsimonious manually-scorable short form, hypothesizing retention of the full PE’s discrimination while optimizing assessment effort and time.

## 2. Methods

### 2.1. Participants

Participants were drawn from the University of Miami Healthy Brain Initiative (HBI), a longitudinal cohort of community-recruited midlife and older adults. Recruitment extended from February 2022 through March 2026, during which the Puppy Escape (PE) story was administered alongside the standard HBI evaluation [12] which included the UDS-3 and subsequently UDS-4. 388 participants completed the PE and then the Craft Story 21 (CS) during two separate study visits, and received a consensus diagnostic classification of cognitively normal (CN, N=153), subjective cognitive impairment (SCI, N=106), or mild cognitive impairment (MCI, N=87); 42 participants who carried a diagnosis of dementia or other cognitive disorder were excluded from primary analyses, resulting in a total of 346 participants for this study. A CN-vs-MCI subsample (n = 240) was used for discrimination and incremental-validity analyses. The full operational rules for diagnostic group assignment, including the cognitive complaint threshold used to assign SCI, follow the cognitive status determination procedure described in the HBI protocol [12]. All study procedures were approved by the University of Miami Institutional Review Board, and all participants provided written informed consent.

### 2.2. Study Design

The present analyses used a cross-sectional, within-participant design in which each participant completed both narrative recall tasks, the neuropsychological battery, and functional and subjective-cognitive measures. PE was administered first alongside plasma collection and other assessments not included in this study, with CS administered on a separate visit 1–3 weeks later alongside the full UDS clinical assessment; intervening neuropsychological testing filled the retention interval between immediate and delayed recall trials for each story. Although task order was not counterbalanced, the minimum one-week between-visit delay is intended to extinguish short-term familiarity and fatigue effects that would otherwise confound fixed-order administration [13,14]. Examiners were not blinded to diagnostic status at the time of administration; PE scoring was performed by the automated pipeline without examiner input on the scored transcripts, and CS scoring was performed by a trained examiner against the standardized NACC UDS-3/4 rubric [6].

### 2.3. Narrative Recall Tasks

#### 2.3.1. Puppy Escape (PE)

The Puppy Escape is a narrative recall task developed as an openly available alternative to existing proprietary narrative recall instruments that restrict clinical and research use. The stimulus is a single paragraph of 124 words describing two friends, a dog’s escape from the home, and a multi-stage search culminating in the dog’s recovery; the story was constructed with a balanced distribution of proper names, spatiotemporal details, action content, and character interactions to permit subscore analysis across semantic categories that total-score paragraph-recall instruments cannot resolve. The full narrative text is reproduced in **Supplementary Files 1** and **2**.

Administration was embedded in a broader automated cognitive battery collecting gaze and speech behavior on a semi-autonomous platform, ensuring consistent conditions across participants. After an instructional period, the narrative was presented bimodally (visual text and audio, male voice) to facilitate encoding and capture cross-modal integration. Immediately following the audiovisual presentation, a blank screen was displayed alongside an audiovisual instruction to “Describe as much of the story as you can remember” (immediate recall, IR). After a filled delay of approximately 15 minutes (determined by preliminary study of 49 adults aged 50+), during which unrelated neuropsychological tasks were administered, the participant was asked to recall the story again without re-presentation of the stimulus (delayed recall, DR). Both recall trials were audio-recorded for subsequent automated scoring.

The PE scoring rubric consists of 57 weighted items, each of which codes the presence or absence of a specific piece of story content in the participant’s recall; items carry differential weights reflecting their informational density, yielding a maximum possible score of 79 points. The 57 items are mapped to six semantic categories (name, location, time, action, description, subject) and to five narrative events spanning the full arc of the story (background, meeting, conversation, search, resolution), such that each item contributes to exactly one category and one event. An immediate-recall total score (range 0–79) and a delayed-recall total score (range 0–79) were computed by summing weighted item scores within each trial, and category and event subscores were computed analogously by summing within subset.

Scoring was performed by an automated pipeline. Audio recordings were transcribed using WhisperX [15] with the Whisper large-v3 model [16], and word-level timestamps were retained for downstream fluency and pausing metrics. Each transcript was scored against the provided PE rubric by the Anthropic Claude API (Sonnet 4.6) using a structured prompt containing the rubric and example scored responses (full prompt and caching specification in the GitHub repository), based on prior work identifying high reliability and similarity to human scorers [17]. The pipeline yielded stable scores across repeated executions on the same input transcript. Scoring prompts were designed for use with both Anthropic and OpenAI models, but can be easily adapted to others.

#### 2.3.2. Craft Story 21 (CS)

The Craft Story 21 is the paragraph-recall task originally introduced by Craft and colleagues [18] and subsequently adopted as the narrative recall component of the NACC UDS-3 neuropsychological battery [6]. CS was administered and scored in strict accordance with the UDS-3 protocol. The story was read aloud once by the examiner, followed immediately by a free-recall trial (immediate recall, IR) and, after a 30-minute filled delay, a second free-recall trial (delayed recall, DR). Each recall trial was scored by a trained examiner under verbatim and paraphrase rules per the UDS-3 protocol. The four resulting scores (IR Verbatim, IR Paraphrase, DR Verbatim, DR Paraphrase) served as the primary CS metrics, and a z-scored composite across all four was computed for sensitivity analyses requiring a single CS summary. CS item-level data was not transcribed; analyses were therefore restricted to the four aggregate scores.

### 2.4. Neuropsychological Battery and Clinical Measures

All participants completed the standard CCBH cognitive and functional battery at the same visit at which the CS was administered (1–3 weeks after PE administration). Global cognition was assessed with the Montreal Cognitive Assessment (MoCA) [19]. Verbal episodic memory was assessed with the Hopkins Verbal Learning Test–Revised (HVLT-R) [20], from which Immediate Recall, Delayed Recall, and Recognition Discrimination scores were retained. Executive function and processing speed were assessed with Trail Making Test Parts A (TMT-A) and B (TMT-B) [21], and the Number Symbol Coding Test [22]. Auditory attention and working memory was assessed with Number Span Forwards and Backwards [1], respectively. Language was assessed with the Multilingual Naming Test (MINT) [23] and animal (semantic) fluency [24]. Visuospatial construction and nonverbal memory were additionally assessed with the Benson Complex Figure copy and delayed recall trials, respectively from the UDS-3/4 battery [6].

Functional status and subjective cognitive decline were characterized with a standardized panel of informant- and self-report instruments, including the Clinical Dementia Rating (CDR) and its sum of boxes (CDR-SB) administered by a trained clinician [25]. Functional independence in instrumental activities of daily living was measured with the Functional Activities Questionnaire (FAQ) [26], and global cognitive and functional impairment was additionally quantified with the Quick Dementia Rating System (QDRS) [27]. Subjective cognitive decline was measured with the Healthy Brain 9 (HB9) subjective cognitive composite [28].

### 2.5. Statistical Analysis

All analyses were conducted in Python 3.10 using statsmodels 0.14.5 [29], scikit-learn 1.7.1 [30], and pingouin 0.5.5 [31]. Multiple comparisons within each pre-specified analysis family were controlled using the Benjamini–Hochberg false discovery rate (FDR) procedure [32], and post-hoc pairwise contrasts following omnibus ANOVA were controlled with Bonferroni correction. Missing data was handled with complete-case analysis within each analytic family. Per-family complete-case n values varied with the predictors included: n = 346 for sample-level descriptive statistics and PE delayed–CS convergent correlations; n = 240 for the CN-vs-MCI ROC and Cohen’s d analyses; n = 233 for the PE–CS DeLong comparison; n = 226 for the hierarchical logistic regression after listwise exclusion across demographics, CS DR Paraphrase, and PE Delayed Total; and n = 326 for the parallel hierarchical linear regressions. Per-family n values are reported in the corresponding Results subsections and table footnotes. Two-tailed tests were used throughout, and alpha was set to .05 for all confirmatory comparisons.

#### 2.5.1. Sample characterization

Between-group differences in demographic and baseline clinical characteristics were evaluated with one-way analysis of variance (ANOVA) for continuous variables and chi-square tests of independence for categorical variables, with diagnostic group (CN, SCI, MCI) as the between-subjects factor. Significant omnibus effects were followed by Bonferroni-corrected pairwise contrasts. Sample characteristics are summarized in **Table 1**.

#### 2.5.2. Convergent validity

Convergent validity between PE and CS was evaluated using Pearson correlations between each PE aggregate score (IR total, DR total) and each of the four CS subscores (IR Verbatim, IR Paraphrase, DR Verbatim, DR Paraphrase). Correlations were computed in the full analytic sample and, separately, within the CN subgroup only, to verify that the association was not driven solely by between-group variance. To evaluate the robustness of the PE–CS association across demographic strata, the same correlations were re-estimated within sex (female, male) and education (< 16, ≥ 16 years) subgroups, with FDR correction applied within each stratum.

#### 2.5.3. Criterion validity

Criterion validity was evaluated by computing Pearson correlations of each PE and CS aggregate with twelve neuropsychological criteria spanning memory (HVLT Immediate, HVLT Delayed, HVLT Recognition, Benson Recall), executive function (TMT-B, Verbal Fluency, Digit Span Backward), processing speed (TMT-A, Number Symbol Coding), language (MINT), visuospatial function (Benson Copy), and global cognition (MoCA Total). Functional criterion correlations were computed in parallel for CDR-SB, FAQ, QDRS, and HB9. Formal comparisons of PE and CS against each shared criterion were carried out with Steiger’s z test for dependent correlations [33]. FDR was applied within three pre-specified families, with Bonferroni correction used in place of FDR for post-hoc pairwise contrasts following omnibus ANOVA.

#### 2.5.4. Group discrimination

CN-vs-MCI group discrimination was quantified in two complementary ways. Effect sizes were computed as Cohen’s d (positive values denoting CN > MCI), both unadjusted and adjusted for age, sex, and education via residualization on the covariate set prior to effect-size computation. Classification performance was evaluated by fitting univariate logistic regression models with each PE or CS score as the sole predictor of CN-vs-MCI status, and computing the receiver operating characteristic area under the curve (ROC-AUC) with 95% confidence intervals by DeLong’s method [34] and by bootstrap resampling (2,000 iterations). Pairwise comparisons between PE DR total and CS DR Verbatim AUCs were carried out with DeLong’s test for correlated AUCs. A combined PE + CS logistic regression model was additionally fit under 10-fold stratified cross-validation, with out-of-fold predicted probabilities used to compute a cross-validated AUC.

#### 2.5.5. Incremental validity

Incremental diagnostic validity of PE beyond CS was assessed with hierarchical logistic regression predicting CN-vs-MCI status. Step 1 included demographic covariates (age, sex, education); Step 2 added the CS DR Paraphrase score, selected a priori based on the established ability of delayed recall to detect MCI; and Step 3 added the PE DR total. Model comparison relied on likelihood-ratio chi-square tests, McFadden pseudo-R², and Akaike information criterion (AIC). Parallel hierarchical linear regressions on continuous cognitive and functional outcomes (MoCA Total, HVLT Delayed, CDR-SB, FAQ Total) followed the same three-step structure with both CS DR Verbatim and DR Paraphrase entered at Step 2 and the four PE delayed-recall features (PE DR total plus three embedding features: narrative similarity, temporal order, and recalled segment proportion) entered together at Step 3. ΔR² and F-change tests were used to evaluate the incremental contribution of the PE feature block beyond demographics and CS at Step 3.

#### 2.5.6. Dimensional profiling

The dimensional structure of MCI-related PE impairment was evaluated with two mixed repeated-measures ANOVAs. In the first model, diagnostic group (CN, SCI, MCI) served as the between-subjects factor and PE semantic category (name, location, time, action, description, subject) served as the six-level within-subjects factor; category scores were standardized within condition prior to analysis to place all six categories on a common scale. In the second model, the same group factor was crossed with a five-level PE event factor (background, meeting, conversation, search, resolution). Models were fit with pingouin’s *mixed_anova* function [31] with Greenhouse–Geisser correction applied when sphericity was violated [35]. Partial eta-squared is reported as the effect-size metric, and significant Group × Subscale interactions were followed by per-subscale CN-vs-MCI Cohen’s d.

#### 2.5.7. Item-level psychometrics

Item-level properties of the PE rubric were summarized using classical test theory indices: item difficulty (proportion of the analytic sample earning the item) and item discrimination (point-biserial correlation of the item score with the total score), computed separately for the IR and DR conditions. Rank-order stability of item difficulties across diagnostic groups was quantified with Spearman’s rho between the CN and MCI difficulty vectors. Differential item functioning was evaluated with chi-square tests comparing CN and MCI item pass rates, with FDR correction across the 57 items.

#### 2.5.8. PE-18 derivation and validation

A shortened, manually-scorable PE form (PE-18) was derived from the full 57-item rubric to minimize effort in manual scoring. A feature selection strategy utilized psychometric ranking based on a composite of item discrimination and CN–MCI DIF gap and constrained by balanced coverage across narrative story segments. The resulting 18 binary items, each scoring 0 or 1, cover all five narrative events and four of the six semantic categories. A PE-18 total score (range 0–18) was computed for each of the IR and DR trials.

All primary analyses were replicated with the PE-18 total in place of the full PE total: convergent validity against CS, criterion validity against the full neuropsychological and functional battery, hierarchical incremental validity, and CN-vs-MCI effect sizes and ROC-AUC with DeLong comparison to CS. Given that the PE-18 item set was derived and validated in the same analytic sample, the resulting effect sizes are interpreted as upper bounds on expected prospective performance, and cross-validation in an independent sample is identified in the Discussion as a required next step.

#### 2.5.9. Sensitivity and moderation analyses

Sensitivity and moderation analyses evaluated whether the primary PE-vs-CS comparisons were stable across demographic strata. The analytic sample was stratified on sex (female, male) and education (< 16, ≥ 16 years), and within each stratum convergent validity, criterion correlations, Cohen’s d for CN-vs-MCI discrimination, and ROC-AUC were recomputed. The primary discrimination and incremental-validity analyses were additionally re-estimated with age, sex, and education entered simultaneously as covariates to characterize covariate-adjusted effect sizes and AUC values.

### 2.6. Data and Code Availability

The Puppy Escape narrative, the 57-item standard scoring rubric, and the 18-item PE-18 rubric are available at no cost via online registration at https://umiamibrainhealth.org/escape-narratives/ under a non-exclusive copyright license jointly granted by the University of Miami and Dr. Michael J Kleiman, and reproduced in **Supplementary Files 1 and 2**. Automated scoring prompts and versioned copies of all files are also located in the GitHub repository: https://github.com/mjkleiman/escape_narratives. The license permits noncommercial use of the materials, including by nonprofit academic and research institutions regardless of funding source, and routine clinical use by healthcare providers, clinics, and hospital systems for direct clinical assessment, diagnosis, and treatment of their patients, whether or not such use is noncommercial. Commercial integration of the materials into a product, platform or service offered to third parties requires separate licensing; inquiries may be directed to the University of Miami Office of Technology Transfer. The narrative text is ©2015 Michael J. Kleiman; the scoring rubrics are ©2026 University of Miami. Participant-level data from the CCBH Healthy Brain Initiative cohort is available to qualified investigators upon reasonable request to the corresponding author and following execution of a data use agreement.

## 3. Results

### 3.1. Sample Characteristics

The total analytic sample comprised 346 participants (CN = 153, SCI = 106, MCI = 87) who had completed both the Puppy Escape (PE) and the Craft Story 21 (CS) during CCBH study visits between February 2022 and March 2026. The three diagnostic groups were well matched on years of education (F(2, 336) = 2.08, p = .126) but differed on age, with MCI participants older than the CN and SCI groups (F(2, 343) = 9.32, p < .001, η² = .052), and on sex distribution, with a lower proportion of female participants in the MCI group (52%) than in the CN and SCI groups (approximately 71 to 73%; χ²(2) = 12.35, p = .002, Cramer’s V = .19). Demographic and baseline clinical characteristics by group are reported in **Table 1**.

### 3.2. Convergent Validity

Puppy Escape total scores correlated moderately with every Craft Story 21 subscore, consistent with both tasks indexing a shared verbal episodic memory construct. In the full analytic sample, PE Immediate Total correlated with CS IR Verbatim, CS IR Paraphrase, CS DR Verbatim, and CS DR Paraphrase at r = .43, .42, .45, and .43, respectively, and PE Delayed Total correlated with the same four CS subscores at r = .43, .42, .44, and .45 (all p < .001, FDR-corrected); the full correlation matrix and 95% CIs are reported in **Table 2**. The association was uniform across scoring rules and recall trials.

Because between-group variance on a clinical-continuum sample can inflate raw convergent validity estimates, the same correlations were recomputed within the CN subgroup alone. Within CN, PE–CS correlations remained significant across all four CS subscores, with r values ranging from .23 to .31 (all FDR-corrected p ≤ .045). The PE–CS correlations remained significant after FDR correction across sex and education strata (female and male; education < 16 and ≥ 16 years), confirming that convergent validity was robust across the major demographic moderators present in the sample.

### 3.3. Criterion Validity

Both PE and CS correlated significantly with every neuropsychological and functional criterion examined, and the magnitude of the PE correlations did not significantly differ from the CS correlations on 10 of 12 neuropsychological comparisons. Pearson correlations (**Table 2**) showed that PE Delayed Total and CS DR Verbatim correlated at comparable magnitudes with global cognition (MoCA Total: PE r = .44, CS r = .42; Steiger p = .64), verbal episodic memory (HVLT Immediate: PE r = .38, CS r = .44, Steiger p = .22; HVLT Delayed: PE r = .48, CS r = .49, Steiger p = .83), executive function (TMT-B: PE r = −.35, CS r = −.29, Steiger p = .22), verbal fluency (PE r = .35, CS r = .36; Steiger p = .61), and language (MINT: PE r = .29, CS r = .21; Steiger p = .14). Two Steiger comparisons reached uncorrected significance, in opposite directions: HVLT Recognition Discrimination favored CS (PE r = .34, CS r = .44; Steiger p = .04), and TMT-A favored PE (PE r = −.31, CS r = −.19; Steiger p = .03). After FDR correction within the 12-criterion family, neither comparison remained significant, and every neuropsychological criterion in the battery yielded an FDR-non-significant Steiger comparison. Both measures correlated significantly with the CDR-SB (PE r = −.35, CS r = −.31; ps < .001) and with the QDRS total score (PE IR r = −.12, p = .013; CS DR Verbatim r = −.18, p < .001), as well as the HB9 (PE r = −.12, p < .05; CS r = −.17, p < .01; Steiger p = .394); on the FAQ, PE reached significance (r = −.12, p < .05) but CS did not (r = −.06), with a non-significant Steiger comparison (p = .377).

### 3.4. Group Discrimination

Score distributions for both narrative recall tasks revealed the same clinical gradient across diagnostic groups: CN and SCI participants produced largely overlapping score distributions on every recall trial, while MCI participants were shifted toward the lower end of the score range. PE Immediate Total (CN 43.8 ± 12.5; SCI 41.2 ± 13.4; MCI 31.5 ± 12.5) and PE Delayed Total (CN 41.9 ± 12.9; SCI 38.3 ± 14.5; MCI 28.8 ± 12.4) distributions (**Figure 1**) placed CN and SCI medians within a narrow band, with the MCI distribution displaced downward and exhibiting a broader spread that extended further into the floor of the score range. The same pattern held for CS Immediate Recall (Verbatim: CN 21.7 ± 5.4, SCI 20.7 ± 5.4, MCI 16.3 ± 5.4; Paraphrase: CN 16.1 ± 3.3, SCI 15.6 ± 3.0, MCI 12.5 ± 3.3) and CS Delayed Recall (Verbatim: CN 19.5 ± 5.1, SCI 18.0 ± 5.8, MCI 12.8 ± 5.8; Paraphrase: CN 15.3 ± 3.5, SCI 14.5 ± 3.6, MCI 10.9 ± 3.9) distributions (**Figure 2**) across both verbatim and paraphrase scoring rules. One-way ANOVAs were significant for every metric (PE Immediate F(2,343)=26.53, PE Delayed F=26.33, CS IR Verbatim F=28.54, CS IR Paraphrase F=38.56, CS DR Verbatim F=41.23, CS DR Paraphrase F=43.11; all p<0.0001), and Bonferroni-corrected pairwise contrasts confirmed that MCI participants scored significantly lower than both CN (all p<0.0001) and SCI (all p<0.0001) on every PE and CS metric, while CN and SCI distributions did not differ significantly on any narrative recall score (PE Immediate p=0.31, PE Delayed p=0.12, CS IR Verbatim p=0.36, CS IR Paraphrase p=0.46, CS DR Verbatim p=0.11, CS DR Paraphrase p=0.23). The convergence between CN and SCI alongside the consistent downward displacement of MCI established that both PE and CS captured the same diagnostic gradient and that the discriminative signal was concentrated at the CN-versus-MCI boundary rather than between the cognitively intact groups.

**Figure 1.**
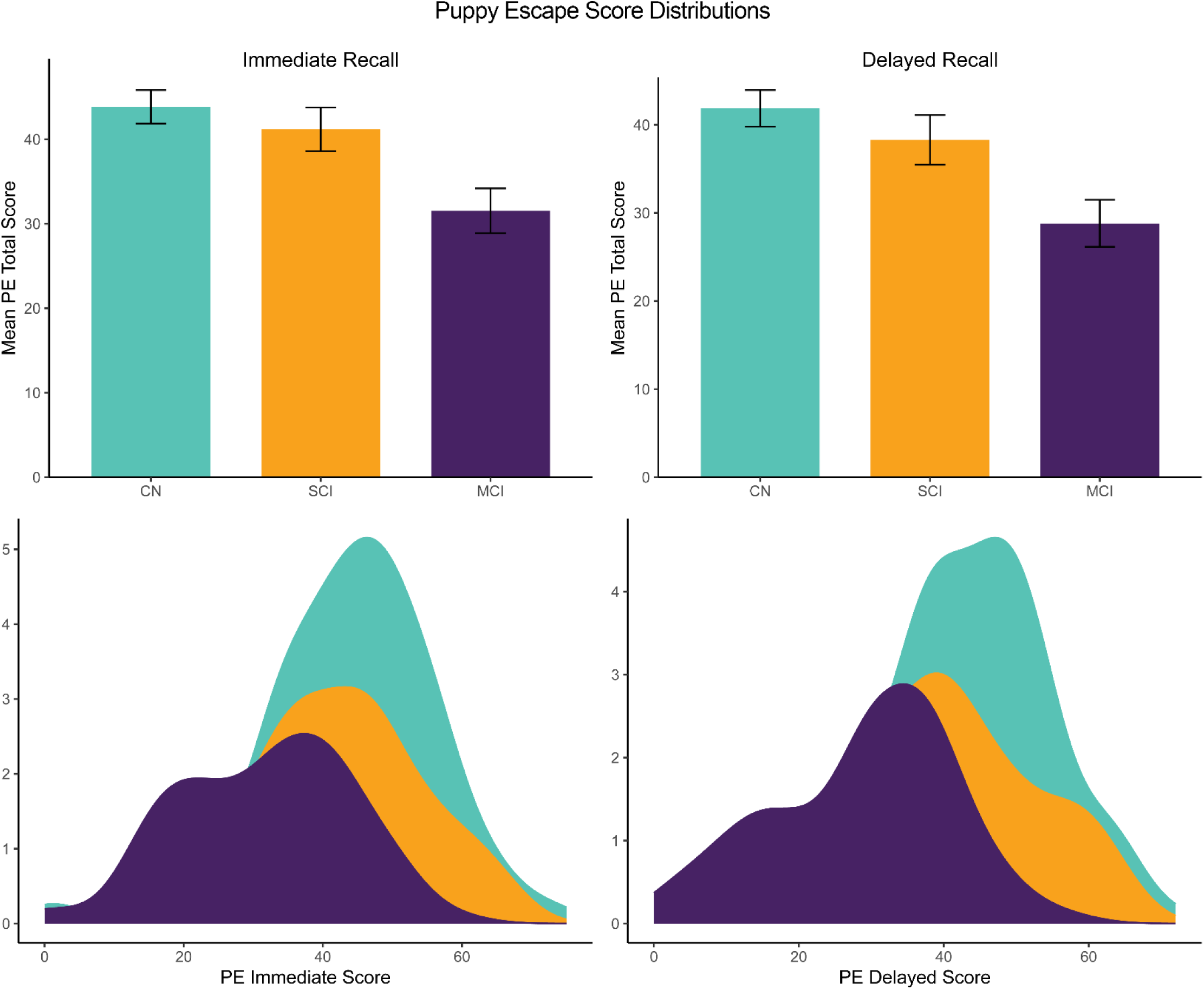
Score distributions of the Puppy Escape (PE) narrative recall task by diagnostic group. The 2×2 panel layout displays Immediate Recall (left column) and Delayed Recall (right column) of the PE Total Score across the three diagnostic groups: cognitively normal (CN, n = 153; teal), subjective cognitive impairment (SCI, n = 106; yellow), and mild cognitive impairment (MCI, n = 87; purple). The top row shows group mean PE Total Scores with error bars representing 95% confidence intervals; the bottom row shows the corresponding kernel density distributions of individual scores. Across both Immediate (CN 43.8 ± 12.5; SCI 41.2 ± 13.4; MCI 31.5 ± 12.5) and Delayed (CN 41.9 ± 12.9; SCI 38.3 ± 14.5; MCI 28.8 ± 12.4) trials, CN and SCI distributions are largely overlapping, whereas the MCI distribution is shifted downward. One-way ANOVAs were significant on both trials (Immediate F(2,343) = 26.53, p < 0.0001; Delayed F(2,343) = 26.33, p < 0.0001). Bonferroni-corrected pairwise contrasts indicated that MCI scored significantly lower than both CN and SCI on Immediate and Delayed Recall (all p < 0.0001), while CN and SCI did not differ (Immediate p = 0.31; Delayed p = 0.12).

**Figure 2.**
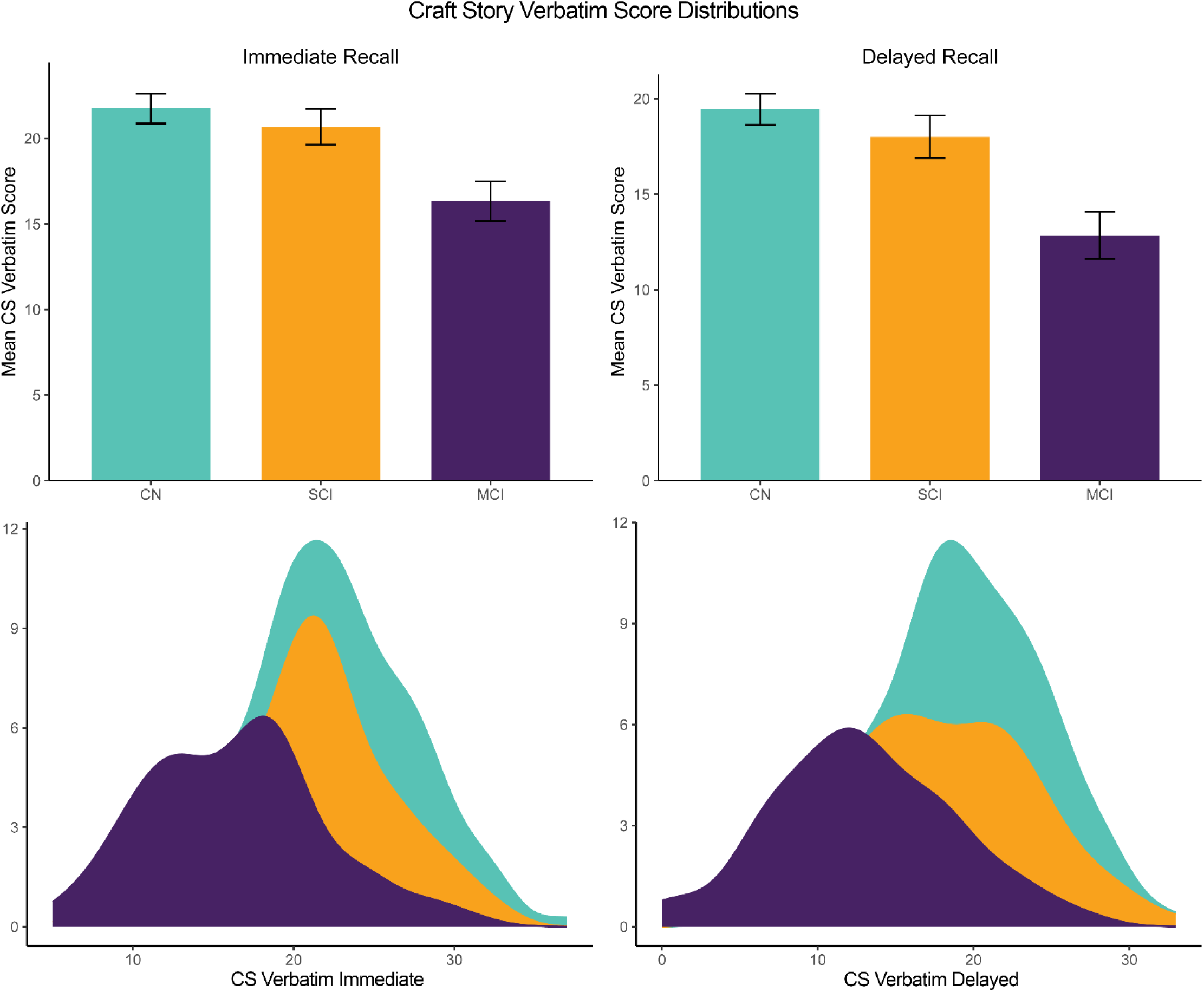
Score distributions of the Craft Story (CS) narrative recall task by diagnostic group. The 2×2 panel layout displays Immediate Recall (left column) and Delayed Recall (right column) of the CS Verbatim score across the three diagnostic groups: CN (n = 153; teal), SCI (n = 106; yellow), and MCI (n = 87; purple). The top row shows group mean CS Verbatim scores with 95% confidence interval error bars; the bottom row shows the corresponding kernel density distributions of individual scores. Across both Immediate (CN 21.7 ± 5.4, SCI 20.7 ± 5.4, MCI 16.3 ± 5.4) and Delayed (CN 19.5 ± 5.1, SCI 18.0 ± 5.8, MCI 12.8 ± 5.8) trials, CN and SCI distributions are largely overlapping, whereas the MCI distribution is displaced downward. One-way ANOVAs were significant on both trials (Immediate Verbatim F(2,343) = 28.54, p < 0.0001; Delayed Verbatim F(2,343) = 41.23, p < 0.0001). Bonferroni-corrected pairwise contrasts indicated that MCI scored significantly lower than both CN and SCI on Immediate and Delayed Recall (all p < 0.0001), while CN and SCI did not differ (Immediate p = 0.36; Delayed p = 0.11). The parallel CS Paraphrase scoring rule showed the same pattern (Immediate F = 38.56; Delayed F = 43.11; all CN/SCI vs MCI contrasts p < 0.0001; CN vs SCI not significant).

Puppy Escape total scores yielded large CN-vs-MCI effect sizes that were not significantly different from the Craft Story 21 at the level of ROC-AUC, with PE Delayed Total exceeding the conventional d = 1.0 threshold. PE Immediate Total and PE Delayed Total produced unadjusted CN-vs-MCI Cohen’s d of 0.99 and 1.03, respectively, and covariate-adjusted d values of 0.90 and 0.98 after residualization on age, sex, and education (**Table 3**). The Craft Story 21 subscores produced numerically larger effect sizes, with CS IR Verbatim at d = 1.00, CS IR Paraphrase at d = 1.13, CS DR Verbatim at d = 1.23 (adjusted d = 1.18), and CS DR Paraphrase at d = 1.22 (adjusted d = 1.17). The numerical gap between PE and CS was concentrated on the delayed recall trial (Δd ≈ 0.20 between PE Delayed Total and CS DR Verbatim), while the immediate recall trial produced essentially equivalent effect sizes against CS IR Verbatim (Δd ≈ 0.01; the gap against CS IR Paraphrase was Δd ≈ 0.14), **Figure 3**.

**Figure 3.**
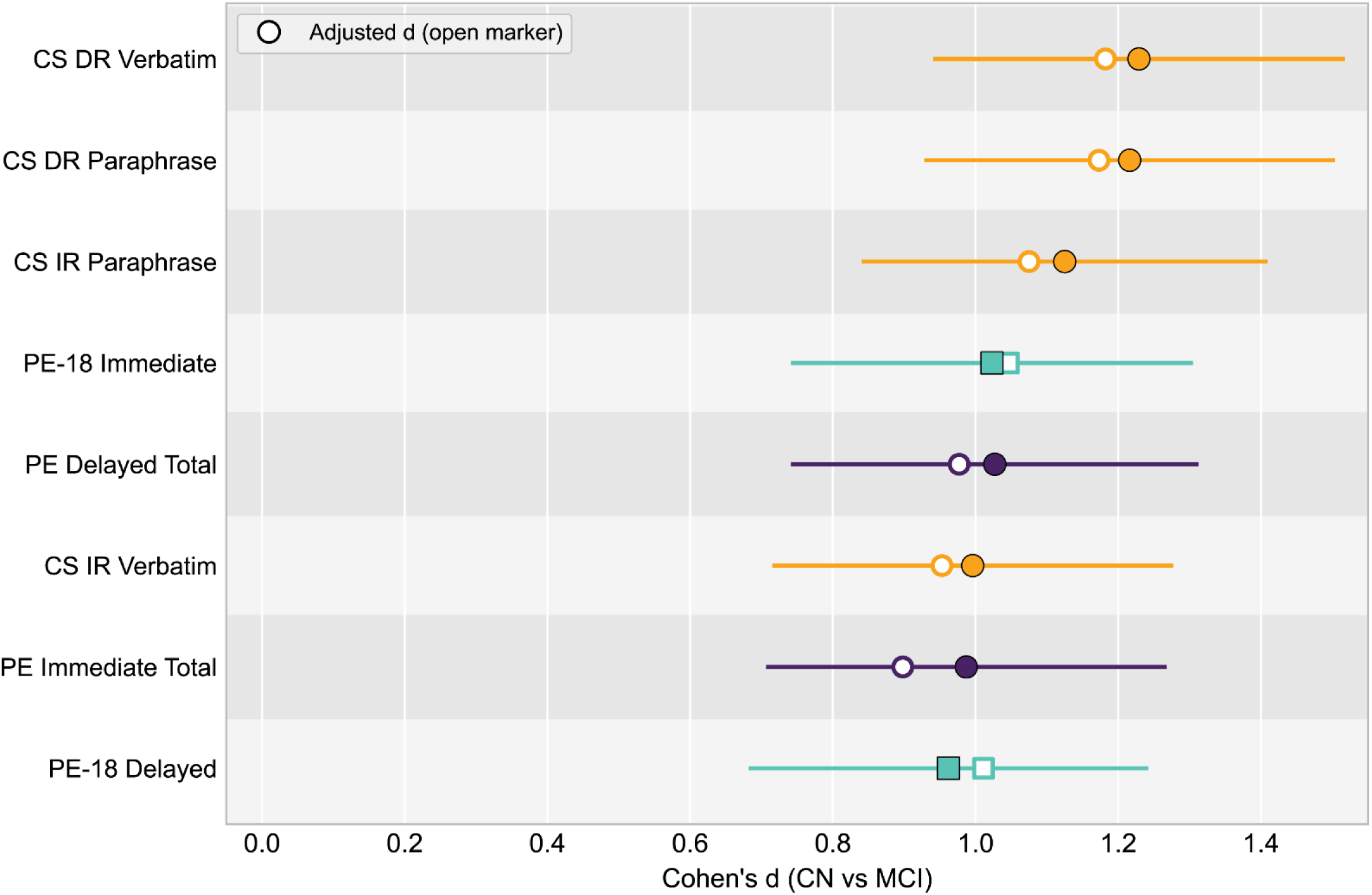
Forest plot of CN-vs-MCI standardized mean differences (Cohen’s *d*) for eight memory measures: Puppy Escape full-scoring totals (PE Immediate, PE Delayed; purple circles), the 18-item subset (PE-18 Immediate, PE-18 Delayed; teal squares), and Craft Story scores (CS IR/DR Verbatim, CS IR/DR Paraphrase; yellow circles). Filled markers show unadjusted *d*; open markers show ANCOVA-adjusted *d* controlling for age, sex, and education. Horizontal lines are 95% parametric confidence intervals around the unadjusted estimate. Larger values indicate greater CN > MCI separation. Effect sizes are computed on complete cases per measure within the n = 346 analytic sample (CN = 153, MCI = 87).

Classification performance, reported in **Table 3**, followed the same pattern: PE Delayed Total achieved an AUC of 0.776 (95% CI [.716, .835]) for CN-vs-MCI discrimination, and CS DR Verbatim achieved an AUC of 0.802 (95% CI [.743, .861]); the CS composite, formed by z-scoring across all four CS scoring rules, reached AUC = 0.806 (95% CI [.748, .863]). The DeLong test for correlated AUCs comparing PE Delayed Total against CS DR Verbatim was non-significant (AUC difference = −0.025, 95% CI [−0.101, 0.050]; z = 0.66, p = .51). A combined PE + CS logistic regression model, evaluated under 10-fold stratified cross-validation, achieved AUC = 0.838 (95% CI [.786, .890]), exceeding the AUC of either story alone and confirming that PE and CS contributed partially non-overlapping information to CN-vs-MCI classification.

### 3.5. Dimensional Profiling

Puppy Escape impairment in MCI was non-uniform across semantic content categories, with a significant Group × Category interaction and a selective pattern of larger CN-vs-MCI effect sizes for location, action, subject, and proper-name content than for descriptive content. The mixed ANOVA modeling the PE subscore with diagnostic group (CN, SCI, MCI) as the between-subjects factor and the six PE semantic categories (name, location, time, action, description, subject) as the within-subjects factor yielded a significant Group × Category interaction (F(10, 1715) = 2.34, p = .010, partial η² = .013), superimposed on a large between-subjects main effect of diagnostic group (F(2, 343) = 23.89, p < .001, partial η² = .122) and a large within-subjects main effect of category (F(5, 1715) = 582.28, p < .001, partial η² = .629). Follow-up pairwise CN-vs-MCI Cohen’s d values, computed within each category, produced the following rank ordering: location (d = 0.85, 95% CI [0.57, 1.12], p < .001), action (d = 0.74, 95% CI [0.47, 1.02], p < .001), subject (d = 0.67, 95% CI [0.40, 0.94], p < .001), name (d = 0.64, 95% CI [0.37, 0.91], p < .001), time (d = 0.52, 95% CI [0.26, 0.79], p < .001), and description (d = 0.21, 95% CI [−0.06, 0.47], p = .379, non-significant), **Figure 4**. The parallel Group × Event mixed ANOVA showed no significant interaction (F(8, 1372) = 1.04, p = .404; per-event Cohen’s d 0.49–0.75), confirming that the selective impairment pattern was specific to semantic content rather than narrative event structure.

**Figure 4.**
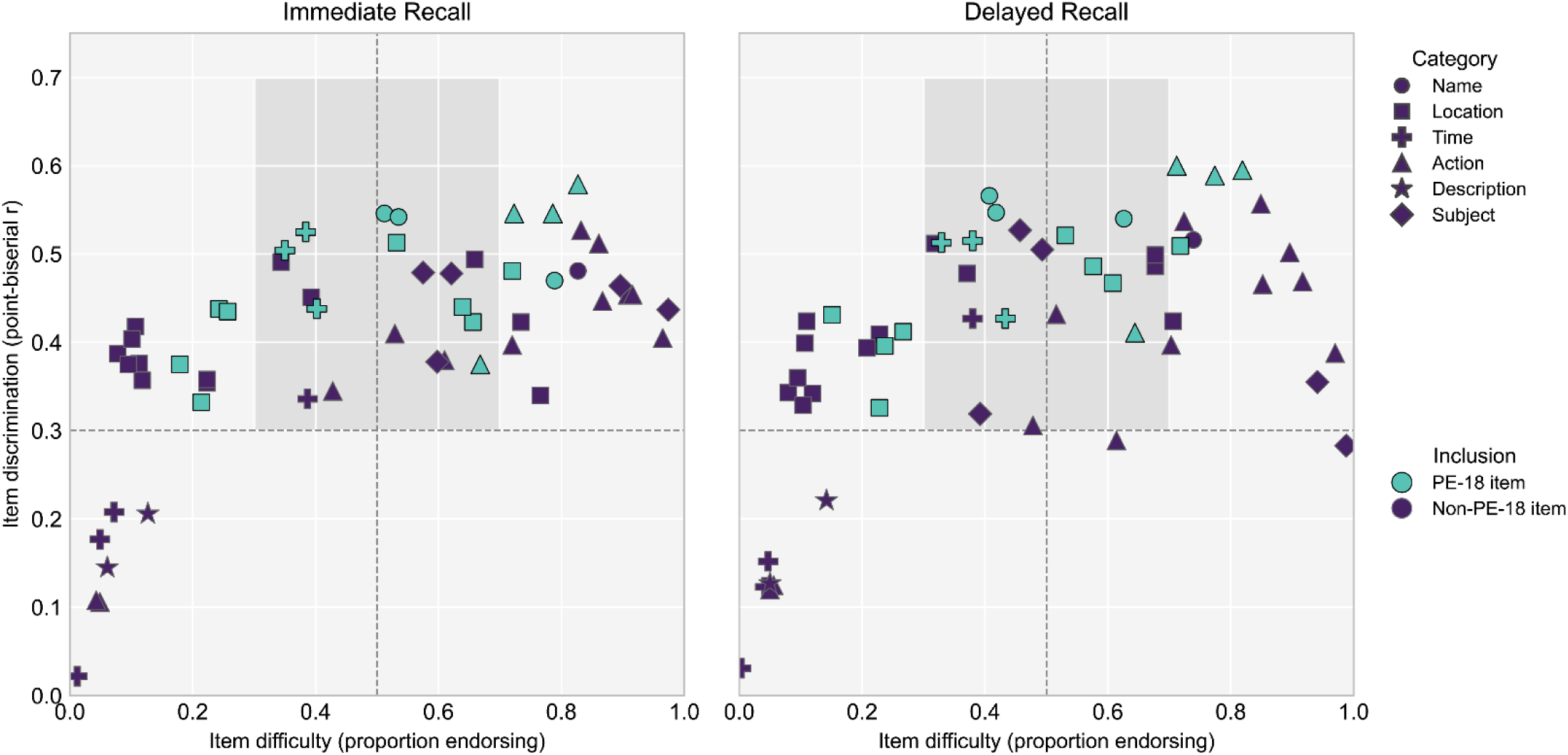
Item-level psychometric properties of all 57 Puppy Escape items in the analytic sample (*n* = 346: CN = 153, SCI = 106, MCI = 87). Left panel: immediate recall (Q000); right panel: delayed recall (Q001). Horizontal axis: item difficulty (proportion of participants endorsing the item). Vertical axis: item discrimination (point-biserial correlation between the binary item score and the recall total). Dashed reference lines mark difficulty = .50 and discrimination = .30; the shaded rectangle delimits the conventional preferred zone (difficulty .30–.70, discrimination .30–.70). Marker shape encodes scoring category: Name (circle), Location (square), Time (cross), Action (triangle), Description (star), Subject (diamond). Fill indicates inclusion in the 18-item short form: teal = PE-18 (*n* = 18), purple = non-retained items (*n* = 39). PE-18 items concentrate inside the preferred zone, while non-retained items predominantly fall outside it (low difficulty or low discrimination).

### 3.6. Incremental Validity

After demographic and Craft Story variance had been accounted for, the Puppy Escape total score added statistically significant predictive variance for CN-vs-MCI classification, and the same incremental pattern held across three of four continuous cognitive and functional outcomes. In the hierarchical logistic regression (n = 226 after listwise exclusion across the demographic, CS, and PE variables entered in the model), a Step 1 model containing age, sex, and education as predictors yielded a McFadden pseudo-R² of .062 and an AIC of 285.8; adding the best-performing Craft Story score (CS DR Paraphrase) at Step 2 produced a substantial improvement (pseudo-R² = .254; AIC = 230.9); and the subsequent addition of PE Delayed Total at Step 3 produced a further significant improvement (pseudo-R² = .308; AIC = 216.9; LR χ²(1) = 16.05, p < .001; ΔR² = +.054). The Step 3 model remained the best-fitting configuration based on both likelihood-ratio comparison and AIC, and PE Delayed Total retained a significant coefficient in the presence of the best CS score and all demographic covariates.

The same incremental pattern replicated in three parallel hierarchical linear regression models predicting continuous cognitive and functional outcomes, with the four PE delayed-recall features (PE DR total plus the three embedding features) entered together at Step 3 beyond demographics and both CS DR scores: MoCA Total (ΔR² = +6.9%; F_change(4, 316) = 7.68, p < .001), HVLT Delayed Recall (ΔR² = +8.0%; F_change(4, 316) = 10.50, p < .001), and CDR Sum of Boxes (ΔR² = +4.0%; F_change(4, 316) = 4.11, p = .003). The FAQ Total model showed no significant incremental contribution from either measure, consistent with the restricted functional impairment range in a sample dominated by CN and early-stage cognitive concerns.

### 3.7. PE-18 Derivation and Validation

An 18-item manually-scorable Puppy Escape short form (PE-18) preserved or modestly exceeded the full PE’s CN-vs-MCI discriminative power, matched it across non-dimensional validation domains, and slightly exceeded the CS in incremental validity. The PE-18 items are reported in **Supplementary File 2** and visually depicted in **Figure 4**. For CN-vs-MCI group discrimination, PE-18 yielded an immediate-recall Cohen’s d of 1.02 and a delayed-recall Cohen’s d of 0.96, retaining the full PE’s effect size (**Table 3**). The corresponding ROC-AUC values were .757 (IR; 95% CI [.694, .821]) and .750 (DR; 95% CI [.688, .812]), and the DeLong comparison of PE-18 Delayed against CS DR Verbatim was non-significant (AUC difference = −0.052, 95% CI [−0.128, 0.025]; z = 1.32, p = .187), mirroring the non-inferior pattern obtained with the full PE.

Across the other validation domains, PE-18 tracked the full PE closely. Convergent validity against CS was slightly attenuated but remained highly significant (r = .38 to .43 across the four CS subscores, all p < .001), and criterion validity against the 12-measure neuropsychological battery yielded non-significant Steiger comparisons on 10 of 12 criteria, with PE-18 numerically outperforming the full PE on TMT-B (r = −.37 vs −.32) and on the Number Symbol Coding Test (r = .37 vs .32). In the hierarchical logistic regression, adding PE-18 Delayed after demographics and the best CS score produced a larger incremental contribution than was obtained with the full PE Delayed Total (ΔR² = +.074, LR χ²(1) = 21.92, p < .001, compared with ΔR² = +.054, LR χ²(1) = 16.05, p < .001 for the full PE), and the parallel hierarchical linear regressions replicated the pattern, with PE-18 adding significant variance beyond CS for MoCA Total (ΔR² = +4.3%, F_change(1, 331) = 19.72, p < .001), HVLT Delayed (ΔR² = +5.4%, F_change(1, 330) = 28.80, p < .001), and CDR Sum of Boxes (ΔR² = +5.0%, F_change(1, 331) = 21.56, p < .001). A combined PE-18 + CS logistic regression model reached AUC = 0.830, approaching the full PE + CS combined model (AUC = 0.838).

The one validation domain in which PE-18 did not replicate the full PE was dimensional profiling. The mixed ANOVA crossing diagnostic group with the PE-18 semantic categories (four of the six original categories are represented in the 18-item subset, with fewer items contributing to each) yielded a non-significant Group × Category interaction (F(6, 1029) = 0.19, p = .980, partial η² = .001), in contrast to the significant interaction obtained with the full PE (F(10, 1715) = 2.34, p = .010, partial η² = .013). PE-18 therefore preserved total-score discrimination and incremental validity but did not retain the content-level selectivity pattern that required the full 57-item automated rubric to detect.

## 4. Discussion

The Puppy Escape matched the Craft Story 21 across every validation domain examined in a research cohort of 346 participants, yielded large CN-vs-MCI effect sizes that exceeded the conventional d = 1.0 threshold and were not statistically distinguishable from CS at the level of ROC-AUC, and captured unique diagnostic variance that CS did not, establishing an openly available narrative recall instrument with psychometric properties on par with the NACC UDS-3 standard. A parsimonious 18-item short form derived from the automated rubric (PE-18) retained the full PE’s total-score discrimination, exceeded the full PE in incremental contribution beyond CS, and can be administered and scored with pen and paper against substantially fewer items than the full CS rubric. Notably, the automated 57-item rubric revealed a selective pattern of MCI impairment on location, action, and proper name content that was not visible at the level of total scores, although was not retained in the manual PE-18 short form.

### 4.1. Assessment Similarity

The PE matched the CS across every validation domain examined in the present cohort. Convergent and criterion validity were statistically similar to CS on 10 of 12 neuropsychological criteria after FDR correction, and CN-vs-MCI discrimination reached d > 1.0 on delayed recall with ROC-AUC not significantly different from CS. The 18-item manually-scorable PE-18 short form provides the most immediately deployable version of the task: it preserves the full PE’s total-score discrimination and slightly exceeds the full PE in incremental variance beyond CS, and can be adopted for routine clinical and research use without per-subject fees, digitization restrictions, or institutional agreements.

### 4.2. The Added Value of Automated Scoring and Dimensional Profiling

The mixed ANOVA crossing diagnostic group with the six PE semantic categories yielded a significant Group × Category interaction, and follow-up CN-vs-MCI effect sizes revealed a selective pattern: location, action, subject, and proper name content were substantially impaired in MCI, while temporal content was impaired to a lesser degree and descriptive content showed no significant group difference. The selective vulnerability of names, locations, and actions, the three content categories carrying the strongest relational load in the PE narrative, is consistent with the contextual-binding deficits documented in early AD [37–39] and suggests that PE captures this deficit through a single-paragraph recall task rather than a dedicated binding paradigm. The relative sparing of descriptive content is consistent with the gist-vs-detail dissociation documented in aging and early AD, in which perceptual event detail declines while narrative gist is comparatively preserved [40–42].

Selective decline in proper name retrieval on Logical Memory–style story recall is associated with β-amyloid burden [43,44] and with broader Alzheimer’s pathology and linguistic markers differentiating MCI from normal aging [45,46]; the fourth-largest CN-vs-MCI effect size on the name category in the present PE rubric extends that observation to MCI in an independent cohort. The category-level resolution required to detect this pattern is unattainable with a total-score paragraph-recall instrument such as the CS.

### 4.3. Incremental Validity and Complementary Information

PE and CS are not perfectly interchangeable: each task captured partially non-overlapping diagnostic variance, with the combined PE + CS model achieving an ROC-AUC of 0.838 and the combined PE-18 + CS model achieving an ROC-AUC of 0.830, each higher than either story alone. In the hierarchical logistic regression, PE Delayed Total added significant variance for CN-vs-MCI discrimination after demographics and the best CS score had been entered, and the same incremental contribution replicated across continuous cognitive and functional outcomes (MoCA, HVLT Delayed Recall, CDR Sum of Boxes). The incremental effect was larger for the 18-item PE-18 short form than for the full 57-item PE total, a pattern suggesting that item reduction via the hybrid psychometric strategy concentrated signal by removing floor and ceiling items without meaningfully reducing construct coverage.

### 4.4. Limitations

The Craft Story 21 retained a numerical effect size advantage on delayed recall (CS DR Verbatim d = 1.23 against PE Delayed Total d = 1.03); although the gap was not statistically significant under the DeLong correlated-ROC test, CS remains the stronger single discriminator in the full-sample analysis, and claims of equivalence should be read as the absence of a statistically detected difference rather than a positive demonstration of empirical equivalence, with a formal non-inferiority claim awaiting a larger sample. The 18-item PE-18 short form was both derived and validated in the same analytic sample, so the reported PE-18 effect sizes are best interpreted as upper bounds on expected prospective performance, and cross-validation in an independent cohort is a required next step before PE-18 is recommended for standalone clinical use. A third constraint is criterion contamination: MCI diagnosis was adjudicated using the same neuropsychological battery that (a) included the Craft Story 21, and (b) subsequently served as criterion-validity targets and outcomes in the hierarchical regressions, such that the effect sizes and criterion correlations reported here are likely inflated relative to those obtainable under a fully independent diagnostic standard.

Several data and design constraints further bound generalizability. The present design is cross-sectional, so longitudinal sensitivity of PE and PE-18 to cognitive decline, test-retest reliability, and the ability to detect conversion from CN or SCI to MCI cannot be evaluated from the present data and represent the most important next steps in the validation program [47,48]. CS item-level data was not transcribed in the HBI export, preventing direct item-level comparison between the two rubrics; CS inter-rater reliability was not assessed in the present sample, although published values are available [6]. Because PE was administered before CS in all participants, CS scores may have been modestly inflated by residual practice effects despite the one-week minimum between-visit interval and the use of distinct narrative stimuli, consistent with practice effects documented across weekly fixed-order neuropsychological administrations even with alternate forms [14]. The MCI group was also less female-skewed than the CN and SCI groups, consistent with documented sex differences in story recall trajectories during preclinical AD [49], and covariate adjustment for sex did not eliminate the primary effects (Section 3.7). Finally, the analytic sample was recruited from a single university-affiliated research center, and the CCBH cohort is self-selected through longitudinal follow-up in the Healthy Brain Initiative; generalizability to community-based samples, to racially and educationally diverse populations [50], to primary care settings, and to international cohorts requires explicit validation in those contexts.

### 4.5. Future Directions

Upcoming studies using this cohort will report (a) computational features unlocked by automated scoring (narrative similarity, temporal organization, discourse coherence, speech production, and machine-learning classifiers), and (b) longitudinal features of the PE narrative, including in-depth examination of biomarkers. Future study will validate the PE and PE-18 short form, as well as an alternative narrative, in an independent cohort.

### 4.6. Conclusions

The Puppy Escape matches the Craft Story 21 across every validation domain examined, providing an openly available narrative recall task with equivalent psychometric properties and partially non-overlapping diagnostic signal. The 18-item manually-scorable PE-18 short form preserves this discriminative power and can be administered and scored with pen and paper. Available at no cost via online registration for noncommercial research use and routine clinical practice, the PE and PE-18 offer a practical path to scalable and equitable cognitive screening in independent research sites, community clinics, federally qualified health centers, and international settings where the cost of proprietary access has been most acute.

## Data Availability

The Puppy Escape narrative, the 57-item standard scoring rubric, the 18-item PE-18 rubric, automated scoring prompts and versioned copies of all files are also located in the GitHub repository: https://github.com/mjkleiman/escape_narratives. Participant-level data from the CCBH Healthy Brain Initiative cohort is available to qualified investigators upon reasonable request to the corresponding author and following execution of a data use agreement.

## ACKNOWLEDGEMENTS

The authors thank the dedicated research participants and their study partners, faculty, staff, postdoctoral fellows, and trainees of the Comprehensive Center for Brain Health at the University of Miami Miller School of Medicine.

## CONSENT STATEMENT

All participants provided written informed consent approved by the University of Miami Institutional Review Board prior to participation in this study.

